# Identification of the Minimal Clinically Important Difference (MCID) for Childhood Autism Rating Scale Second Edition (CARS2) in children with ASD

**DOI:** 10.64898/2026.08.31.26361823

**Authors:** Andrey Vyshedskiy, Lucas Ernesto Pavoski Poloni, Andriane Schmiedel Fucks, Edward Khokhlovich, Elielton Fucks, Andressa Schmiedel

## Abstract

**Purpose:** In clinical trials, treatment efficacy is commonly assessed by comparing control and treatment groups. However, in large samples, even small and clinically trivial differences may achieve statistical significance. Accordingly, the Minimal Clinically Important Difference (MCID) is used as a threshold to determine whether statistically-significant effects are also clinically meaningful to patients. The objective of this study was to estimate the MCID for the *Childhood Autism Rating Scale Second-Edition* (CARS2) using the Patient Impression of Change (PIC) as an external anchor.

**Methods:** Single-item PICs are not well suited to characterizing improvement in a multifaceted disorder such as ASD. Accordingly, the 77-item *Autism Treatment Evaluation Checklist* (ATEC) was used as a multi-item PIC. CARS2 and ATEC were administered concurrently to 62 children with ASD, aged 1.8–7.9 years, with assessments conducted six months apart.

**Results:** The correlation between changes in CARS2 and ATEC total scores was 0.41–0.44 (p<0.0001), supporting the use of ATEC as an anchor measure. Two anchor-based methods yielded MCIDs of 2.39–2.82 CARS2 points. Two distribution-based methods produced MCIDs of 1.33–3.32.

**Conclusions:** Taken together, these approaches suggest that a between-group difference of 2.6 CARS2 points (the midpoint of the anchor-based estimates) may serve as the MCID in children with ASD.

## Introduction

In randomized controlled trials (RCTs), treatment effectiveness is typically evaluated by comparing outcomes between groups (group-level analyses). However, even trivial differences may reach statistical significance in large samples. To address this limitation, researchers use the Minimal Clinically Important Difference (MCID) to determine whether statistically significant changes are also meaningful to patients and clinicians. Originally, MCID was defined as “*The smallest difference in score in the domain of interest which patients perceive as beneficial and which would mandate, in the absence of troublesome side effects and excessive cost, a change in the patient’s management*” (Jaeschke et al., 1989). MCID is commonly estimated as the mean improvement in an outcome measure among patients who report the smallest clinically meaningful change.

In addition to group-level analyses in RCTs, comparing the proportion of individuals who improve (termed “responders”) provides important complementary information. However, the MCID does not account for the measurement error associated with individual change scores and therefore is generally lower than the magnitude of change required to establish true improvement at the individual level. Consequently, using MCID to classify responders can lead to overestimation of treatment effects, with too many individuals incorrectly identified as showing an improvement (Hays & Peipert, 2021). A fundamental criterion for defining a responder is that the observed improvement exceeds measurement error, indicating a true change. This is typically assessed using the Minimal Detectable Change, defined as the smallest change in score that can be interpreted as real with 95% confidence (MDC95). MDC95 increases with the amount of measurement “noise” in the outcome measure and is estimated from the standard error of measurement (SEM), which is derived from test–retest reliability.

In summary, MCID and MDC95 serve distinct and complementary purposes: MCID is used to interpret differences between group averages, whereas MDC95 is used to identify individual responders to treatment. MCID is typically smaller than MDC95 because random measurement error tends to average out in large samples, allowing even modest between-group differences to reflect clinically meaningful population-level effects. In contrast, detecting true change in individuals requires a higher threshold (i.e., MDC95) to account for measurement errors.

Consequently, applying MDC95 to group-level comparisons may be overly conservative, while using MCID to classify individual responders can inflate estimates of improvement by including individuals whose observed changes do not exceed measurement error.

This distinction between MCID and MDC95 is well established and has been empirically examined across many health conditions and measurement instruments (Chaidaroon et al., 2023; Hays & Peipert, 2021). Among cognitive assessments, for example, the Montreal Cognitive Assessment (MoCA) is widely regarded as a standard screening tool. Thousands of studies have used the MoCA to track cognitive changes in patients with dementia and stroke. Consequently, MCID and MDC95 were independently established for the MoCA across various clinical conditions. For example, in dementia populations, the MCID is typically estimated at approximately 2 MoCA points (Krishnan et al., 2017), whereas the MDC95 is estimated at about 4 MoCA points (Feeney et al., 2016). In practical terms, a between-group difference of ∼2 MoCA points in an RCT indicates a clinically meaningful treatment effect. However, for an individual patient, a change of ∼4 MoCA points or more is required to conclude that a true change in clinical status has occurred (with 95% confidence), rather than reflecting random measurement variability. A similar pattern is observed in post-stroke populations, where the discrepancy between MCID and MDC95 is even larger. The MCID for stroke patients is typically estimated at approximately 1.6–2.0 MoCA points, whereas the MDC95 is estimated at about 5.1 MoCA points (Lindvall et al., 2024).

In the field of autism, the *Childhood Autism Rating Scale, Second Edition* (CARS2) (Schopler et al., 2002) has served as a widely used standardized assessment tool (Supplementary Table 1). However, neither its MCID nor MDC95 has been empirically established. Instead, several studies have relied on variably defined thresholds for responder analyses (an individual-level construct), and, to our knowledge, no study has attempted to determine an MCID (a group-level construct). Compounding this issue, Jurek et al. (Jurek et al., 2022) mischaracterized their responder-analysis threshold (an individual-level construct) as “*minimum clinically relevant change*,” a proprietary term later misinterpreted as MCID. Given its subsequent use as a proxy for MCID, this threshold warrants careful scrutiny.

### Examination of the study by Jurek et al. and the prior literature which it relied upon

Jurek et al. (Jurek et al., 2022) sought to establish a threshold for responder analysis. As they note, “*Efficacy claims of an intervention require not only statistical significance but also clinical meaningfulness. One proposed approach to address this question is a responder analysis, in which a continuous primary efficacy measure is dichotomized into ‘responders’ and ‘non-responders’.*” As discussed above, a responder threshold is most appropriately addressed using the MDC95 derived from test-retest reliability data. However, Jurek et al. did not rely on test-retest data. Instead, they employed the Sheffield Elicitation Framework (SHELF), aggregating thresholds reported in prior randomized controlled trials (RCTs).

Jurek et al. cite five references to support their elicitation process (Jurek et al., 2022). The researchers describe compiling an evidence dossier summarizing CARS2 score changes deemed clinically relevant in prior trials: *“An evidence dossier was circulated prior to the elicitation workshop (Supplementary Material, S2). This dossier provided summaries of published evidence on CARS2-2 score changes that were interpreted as being clinically relevant in clinical trials [9, 14–17].”*

**1. Reference 9 (regulatory guideline).** This reference is a regulatory guideline (EMA, 2020), containing neither responder analyses nor threshold estimates, and therefore does not contribute relevant empirical evidence. Its inclusion in the elicitation process is unclear.
**2. Reference 14 (Chez et al.)** This RCT of secretin in ASD employed a responder analysis using a 6-point improvement on CARS2 (Chez et al., 2000). Importantly, this threshold was not intended as an MCID for group-level interpretation, but solely as a criterion for identifying individual responders.
**3. Reference 15 (Coniglio et al.)** In another secretin trial (Coniglio et al., 2001), a responder threshold of 4.07 points was used. As in the previous study, this value was selected for responder analysis and was not proposed as an MCID for group-level interpretation.
**4. Reference 16 (Lemonnier et al.)** In a trial of bumetanide (Lemonnier et al., 2017), responders were defined as individuals with ≥6-point reductions in CARS2: “*patients who achieved a reduction of six points or more on the CARS2 between screening and Day 90*.” However, Jurek et al. instead used a 4-point threshold (see Table 1 in Jurek et al. (Jurek et al., 2022)), possibly derived from a sample size calculation rather than a responder definition. The basis for this substitution is unclear.
**5. Reference 17 (Nagaraj et al.)** In a randomized, placebo-controlled trial of risperidone (Nagaraj et al., 2006), responder status was defined as a ≥20% improvement from baseline.

**Table 1.** Minimal Clinically Important Difference (MCID) estimates for the *Childhood Autism Rating Scale–Second Edition* (CARS2) using anchor-based and distribution-based approaches. The parent-reported *Autism Treatment Evaluation Checklist* (ATEC) was utilized as the Patient Impression of Change (PIC) anchor for anchor-based estimates.

| Method of MCID calculation | N observations | MCID (CARS2 points) | 95% CI | P-Value |
| --- | --- | --- | --- | --- |
| Anchor-based approach: 10–40 point improvement in ATEC total score | 26 | 2.39 | 0.51 – 4.26 | 0.019 |
| Anchor-based approach: 10–20% improvement in ATEC total score relative to baseline | 31 | 2.82 | 1.54 – 4.10 | 0.0002 |
| Distribution-based approach: 0.2–0.5 SD of baseline CARS2 score variability | 62 | 1.33 – 3.32 | N/A | N/A |
| Distribution-based approach: Standard Error of Measurement (SEM) | 62 | 1.8 | N/A | N/A |

In summary, Jurek et al. stated that their goal was to derive a threshold for responder analysis (an individual-level construct) and, accordingly, relied on three studies that explicitly employed responder thresholds for this purpose (the rationale for adopting a different threshold from the fourth study by Lemonnier et al. (Lemonnier et al., 2017) remains unclear). Crucially, Jurek et al. neither aimed to estimate a group-level construct, the MCID, nor relied on studies that had estimated an MCID.

Unfortunately, the responder threshold identified by Jurek et al. was later misinterpreted as an MCID, largely because Jurek et al. referred to it using a proprietary term “*minimum clinically relevant change*,” which is easily conflated with MCID and was subsequently treated as such in later studies (Fradkin et al., 2024; Holeva et al., 2024).

No study has yet established an MCID for CARS2, an important parameter for interpreting the clinical meaningfulness of between-group differences in RCTs involving individuals with ASD. The present study addresses this gap by empirically deriving an MCID for CARS2, thereby providing an appropriate benchmark for group-level comparative analyses in ASD.

## Methods

The study protocol received Institutional Review Board (IRB) approval from Centro Universitário Dinâmica das Cataratas (Foz do Iguaçu, Brazil). The study was conducted in accordance with the ethical standards of the American Psychological Association. Written informed consent was obtained from the parents of all participants.

### Participants

Children diagnosed with ASD and attending the Soulmare - Clínica de Autismo (previously known as Somare therapeutic and educational clinic) in Foz do Iguaçu, Brazil, were recruited for the study (N = 62). The mean age was 4.5 ± 1.3 (range: 1.8 to 7.9 years); 76% were males. Participants were evaluated longitudinally at 6-month intervals using the clinician-administered *Childhood Autism Rating Scale Second Edition* (CARS2), a 15-item instrument with total scores ranging from 15 to 60 (Table S1) (Schopler et al., 2002). The CARS2 has been validated in multiple studies (Moon et al., 2019; Stevanovic et al., 2021), and the Brazilian version used in this study has also been validated in several studies (Backes et al., 2014; Pereira et al., 2008; Rapin & Goldman, 2008). The mean baseline CARS2 was 29.4 ± 6.6 (range: 18 to 48.5).

### Statistical analysis

#### Single-item versus multi-item Patient Impression of Change (PIC)

The Minimal Clinically Important Difference (MCID) is usually estimated using the Patient Impression of Change (PIC) as an external anchor. The PIC is intended to capture the patient’s holistic assessment of treatment benefit by integrating changes across symptoms, function, and quality of life. Because it reflects the patient’s own perception of improvement, the PIC provides a patient-centered criterion for determining whether observed changes in clinical outcome measures are meaningful.

The most widely used PICs employ a single Likert-scale item (e.g., “improved,” “slightly improved,” “no change,” “slightly worsened,” “worsened”). Although the single-item PIC is simple to administer and captures the patient’s overall perception of treatment benefit, it has several important inherent limitations compared with a comprehensive multi-item PIC (Rampakakis et al., 2015; Scott & McCracken, 2015):

1. The most fundamental limitation of a single-item PIC is its low resolution, making it relatively insensitive to small but clinically meaningful changes. In contrast, a PIC derived from dozens of items yields substantially greater measurement precision and is better able to detect gradual improvement over time.
2. Single-item PICs are also more susceptible to measurement error because they rely on a single global judgment. Multi-item PICs reduce random error by averaging responses across numerous questions, resulting in substantially higher reliability and responsiveness. Consequently, multi-item PICs generally provide greater statistical power to detect treatment effects and permit more precise estimation of change.
3. An additional advantage of a multi-item PIC is that it can reduce the limitations of human memory by systematically prompting evaluation across multiple specific dimensions. In a single-item PIC, patients or caregivers must spontaneously recall the most salient changes since treatment initiation, and less noticeable but meaningful improvements may be overlooked. A multi-item PIC serves as a structured reminder of relevant developmental domains and specific behaviors, allowing respondents to recognize changes that may not have been immediately apparent. By sampling across multiple specific dimensions, the multi-item PIC provides a more complete representation of developmental progress and reduces the influence of selective recall. For these reasons, multi-item PICs are generally considered superior for quantifying treatment efficacy (Baker et al., 2024; Challoumas et al., 2026; Charlton et al., 2025; Zhang et al., 2023a).

Furthermore, a single-item PIC may be particularly poorly suited to assessing change in children with ASD. In a separate pilot study (unreleased findings), we evaluated the utility of a single-item PIC for assessing changes in language comprehension in a cohort of 13,251 participants with ASD aged 2–8 years (mean = 4.7 ± 1.5), with evaluations conducted approximately four months apart. The distribution of parent responses was highly skewed toward reporting an improvement: 80% of parents reported that their child’s language comprehension had improved, whereas 16% reported no change, leaving only 4% of responses indicating a decline. This restricted distribution reduced the correlation between change in the single-item PIC and change in a language measure to approximately 0.1, limiting its suitability as an external anchor for MCID estimation.

A single-item PIC is well suited to assessing patient-perceived change when responses are reasonably distributed across all available categories. For example, it is useful for evaluating changes in pain following surgery, changes in cognitive function following an intervention, improvements in fibromyalgia, or changes in sleep quality. Conversely, our findings suggest that a single-item PIC may not be suitable for evaluating parent-perception change in young children with ASD, because responses are overwhelmingly concentrated in the “improved” category. Whether this skew reflects genuine developmental gains, overly optimistic reporting, or the multifaceted nature of ASD (whereby some facets are always improving), the resulting ceiling effect limits the ability of a single-item PIC to discriminate among different degrees of improvement and to serve as an external anchor for estimating MCID.

Accordingly, in this study, we sought to use a multi-item PIC. Rather than developing a proprietary multi-item PIC, we utilized the freely available parent-reported assessment *Autism Treatment Evaluation Checklist* (ATEC) (Rimland & Edelson, 1999) (Supplementary Table 2). The ATEC has been validated in multiple studies (AF et al., 2018; Al Backer, 2016; de Oliveira et al., 2025; Elvitigala et al., 2024; Freire et al., 2018; Geier et al., 2013; Ghosh et al., 2015; Magiati et al., 2011; Mahapatra, Khokhlovich, et al., 2018; Mahapatra, Vyshedsky, et al., 2018; Netson et al., 2024; Nowak et al., 2018; Paveenakiattikhun et al., 2025; Sobaniec et al., 2017; Starling, 2017), and the Brazilian version used in this study has also been validated in several studies (Freire et al., 2018; Montenegro et al., 2021; Santos et al., 2025; Silva Junior et al., 2024). The ATEC consists of 77 items. Thus, rather than relying on a single-item PIC rating (e.g., “[my child] slightly improved”), the ATEC captures parent-perceived improvement across 77 specific dimensions, including language, cognition, sociability, and health-related behaviors. The total ATEC score has been used as PIC (range from 0 to 179). ATEC was completed concurrently with CARS2 assessments. The mean baseline ATEC was 55.0 ± 24.3 (range: 10 to 115). All 168 paired CARS2/ATEC observations were pooled for analysis.

#### Anchor-based method for MCID calculations

With single-item PICs (e.g., “improved,” “slightly improved,” “no change,” “slightly worsened,” “worsened”), the MCID is commonly estimated using an anchor-based approach in which the mean change in the outcome measure among patients or caregivers reporting a minimal but meaningful improvement—typically categorized as “slightly improved”—is used as the estimate of the MCID (Butler et al., 2022). In contrast, a multi-item PIC captures improvement across multiple domains and therefore does not yield a single categorical anchor corresponding to “slightly improved.” Accordingly, the MCID is typically estimated for a multi-item PIC by using a meaningful change in the PIC total score as the external anchor (Fijen et al., 2023). This change can be calculated either as an absolute difference between baseline and follow-up scores or as a relative percentage change from each participant’s baseline score (Zhang et al., 2023a).

Setting an anchor in terms of an absolute number of points is simpler but can be misleading, especially for a cohort with highly variable scores across children. For example, in a child with a very high baseline ATEC score (e.g., 150 points), a 10% reduction (15 points) may reasonably reflect a minimal but meaningful improvement. In contrast, applying the same absolute anchor (15 points) to a child with a lower baseline score (e.g., 30 points) would correspond to a 50% reduction in symptoms, reflecting a major clinical improvement rather than a minimal clinically important difference (Zhang et al., 2023a).

A commonly used alternative is to define an anchor as a relative improvement from each participant’s baseline score, with a 10–15% or 10–20% improvement threshold widely used as a pragmatic benchmark across a variety of clinical outcome measures (Lin et al., 2010; Wu et al., 2019; Zhang et al., 2023a). By defining an anchor as a percentage of baseline severity, the threshold scales proportionally to each child’s symptom burden. Thus, a child with a baseline ATEC score of 150 would require 22-point reduction to meet the 15% improvement threshold, whereas a child with a baseline score of 40 would require 6-point reduction to meet the 15% improvement threshold.

Therefore, the present study estimated the MCID using both types of anchors, defined by absolute and relative improvement in the PIC score. For the absolute-change approach, the MCID was estimated as the mean change in CARS2 score among participants with a 10–40 point improvement in ATEC total score. For the relative-change approach, the MCID was estimated as the mean change in CARS2 score among participants with a 10–20% improvement in ATEC total score from baseline.

Because pooled longitudinal data was utilized, the MCID was calculated using an intercept-only linear mixed-effects model with participant ID set as a random effect to account for within-subject correlations.

#### Distribution-based methods for MCID confirmation

Two distribution-based methods have been used to confirm MCID. The first, the Standard Deviation (SD) method, uses baseline variability within the study population to assess clinical meaningfulness. In this approach, 0.5 SD of the baseline score is commonly interpreted as representing a clinically important change (Norman et al., 2003), whereas 0.2 SD is considered the lower threshold for detecting a minimal meaningful change (Hawkes et al., 2004; Lin et al., 2010; Lindvall et al., 2024). This 0.2–0.5 SD range is frequently used in clinical research, particularly when an anchor-based approach reflecting patient-perceived improvement is unavailable.

Additionally, MCID was estimated using the Standard Error of Measurement (SEM) to account for the inherent “noise” or measurement error of CARS2 (Revicki et al., 2008). The SEM was derived using the formula SEM = SD × √(1 - *r*), where SD denotes the baseline standard deviation and *r* represents test-retest reliability coefficient. This approach provides a precision-based benchmark that reflects the reliability of the CARS2 within the study population, ensuring that the identified change exceeds measurement error and is statistically robust.

## Results

A total of 62 children diagnosed with ASD were enrolled in the study. The mean age of participants was 4.5 ± 1.3 years (range: 1.8–7.9 years). The Minimal Clinically Important Difference (MCID) for the *Childhood Autism Rating Scale Second Edition* (CARS2) was estimated using two complementary approaches: anchor-based and distribution-based.

An anchor-based approach uses the Patient Impression of Change (PIC) to capture a patient’s or caregiver’s overall perception of treatment benefit. Although traditional PICs typically rely on a single Likert-scale item (e.g., “improved,” “slightly improved,” “no change,” “slightly worsened,” “worsened”), such measures have limited resolution, are more susceptible to measurement error, are vulnerable to selective recall, and may be poorly suited for children with ASD because caregiver responses are overwhelmingly skewed toward reports of improvement (see Methods).

In contrast, multi-item PICs provide greater precision, reliability, responsiveness, and statistical power by integrating changes across multiple domains. Accordingly, this study utilized the parent-reported *Autism Treatment Evaluation Checklist* (ATEC) (Rimland & Edelson, 1999) as a multi-item PIC. The ATEC is a validated 77-item instrument that captures perceived changes across multiple developmental domains, including language, cognition, sociability, and health-related behaviors. The total ATEC score (range 0–179) was used as the PIC measure and was collected concurrently with CARS2 assessments. Participants were assessed every six months. All paired CARS2/ATEC observations (N=168) were pooled for MCID analysis.

For single-item PICs, MCID is commonly estimated using an anchor-based approach in which the mean change in an outcome measure among patients or caregivers reporting the smallest clinically meaningful improvement (“slightly improved”) serves as the anchor. Because a multi-item PIC does not yield a single categorical anchor, MCID must instead be estimated using a predefined meaningful change in total PIC score. This change can be defined as either an absolute point difference or a relative percentage change from baseline. In this study, MCID was estimated using both absolute and relative ATEC improvement as anchors. The absolute-change approach estimated the MCID as the mean CARS2 change among participants with a 10- to 40-point improvement in ATEC total score, whereas the relative-change approach estimated the MCID as the mean CARS2 change among participants with a 10 to 20% improvement in ATEC total score from each participant’s baseline (Lin et al., 2010; Wu et al., 2019).

The first step in anchor-based MCID derivation is to establish the correlation between the measure of interest (CARS2) and the PIC. Current recommendations suggest a minimum correlation coefficient of 0.3 for establishing the suitability of an external anchor (Zhang et al., 2023b).

Figure 1A shows changes in CARS2 scores over the 6-month interval in relation to corresponding absolute changes in ATEC total scores. Because the dataset included nested longitudinal data (multiple observations per participant), traditional Pearson or Spearman correlations were inappropriate because they violate the assumption of independence. A repeated measures correlation (rmcorr (Bakdash & Marusich, 2017)) between changes in CARS2 and absolute changes in ATEC was 0.41 (p<0.0001), exceeding the recommended minimum threshold of 0.3 for establishing the suitability of an external anchor (Zhang et al., 2023b).

**Figure 1.**
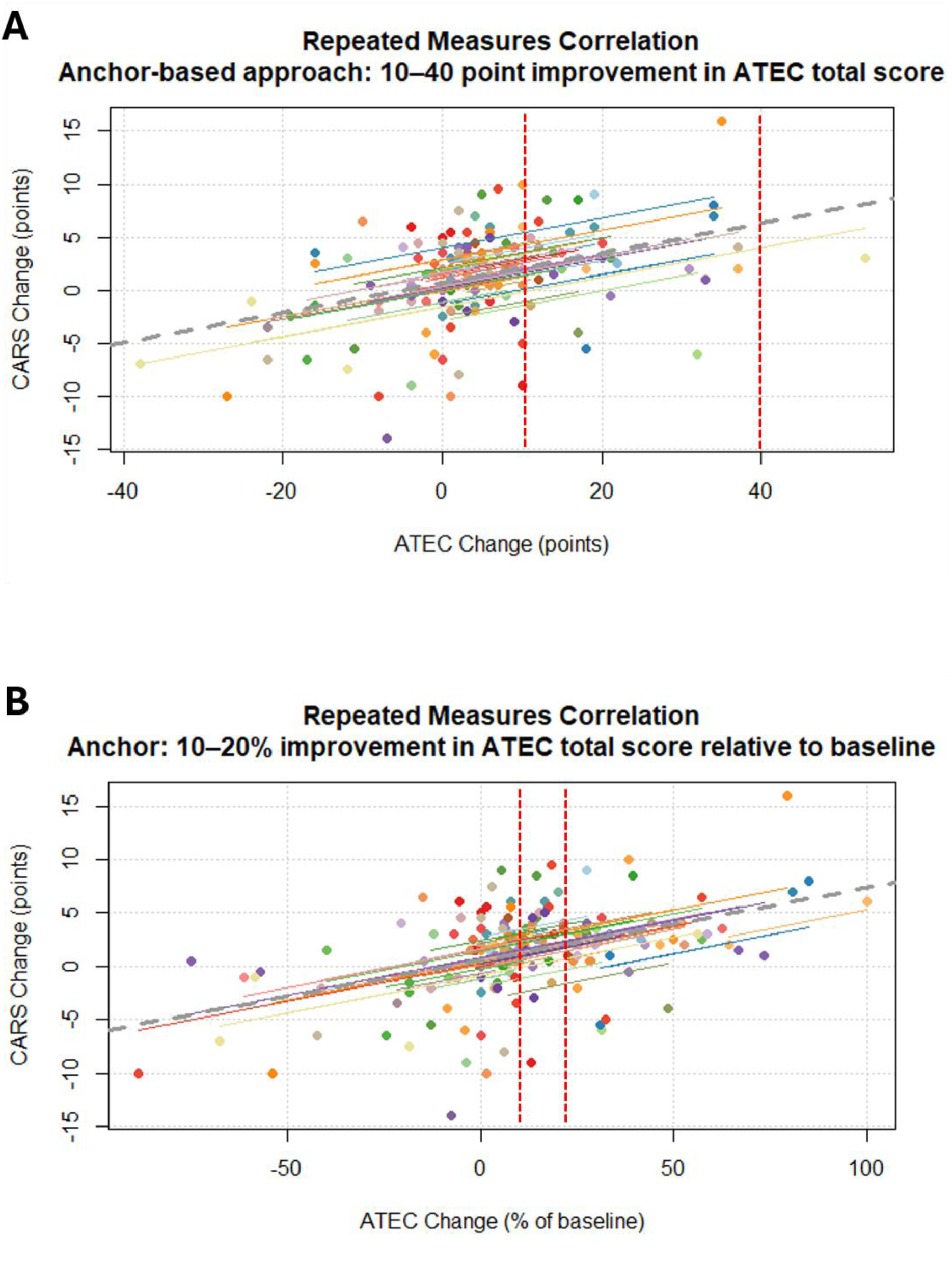
Repeated measures correlation between ATEC and CARS2 improvement scores. Positive values on both axes denote clinical improvement (a reduction in symptom severity). Each distinct color represents an individual participant, with data points indicating their multiple evaluations over time. The colored parallel lines represent the repeated measures correlation fit demonstrating the common within-subject trend while controlling for between-subject baseline differences. For comparison, the dashed black line represents the overall simple regression line, which ignores the nested, longitudinal structure of the data. (A) The scatterplot illustrates the within-subject association between the change in CARS2 (measured in raw score points) and the corresponding change in ATEC total score (r = 0.41, p < 0.0001). Red vertical dashed lines indicate the range 10–40 point improvement in ATEC total score. (B) The scatterplot illustrates the within-subject association between the change in CARS2 (measured in raw score points) and the corresponding change in ATEC total score expressed as % of baseline (r = 0.44, p < 0.0001). Red vertical dashed lines indicate the range 10–20% improvement in ATEC total score relative to each participant’s baseline score.

Figure 1B shows changes in CARS2 scores over the 6-month interval in relation to corresponding relative changes in ATEC total scores from each participant’s baseline. The repeated measures correlation was 0.44 (p<0.0001), again exceeding the recommended minimum threshold of 0.3 for establishing the suitability of an external anchor.

To estimate MCID based on the absolute change in ATEC total score, the mean change in CARS2 score was calculated among participants who exhibited a 10- to 40-point improvement in ATEC total score (N = 26). Because longitudinal observations were pooled, MCID estimates were derived using an intercept-only linear mixed-effects model with participant ID included as a random effect to account for within-subject correlations. The estimated MCID for the CARS2 was 2.39 points (95% CI: 0.51 – 4.26; p = 0.019) (Table 1). Sensitivity analysis of the MCID based on absolute change in ATEC total score demonstrated excellent stability, with estimates ranging from 1.92 to 2.87 points (Table 2).

**Table 2.** Sensitivity analysis for an anchor based on the absolute change in ATEC total score approach.

| Absolute improvement in ATEC total score | N observations | MCID (CARS2 points) | 95% CI | P-Value |
| --- | --- | --- | --- | --- |
| 10–40 points | 26 | 2.39 | 0.51 – 4.26 | 0.019 |
| 9–39 points | 28 | 2.02 | 0.30 – 3.74 | 0.027 |
| 8–38 points | 29 | 1.92 | 0.32 – 3.50 | 0.024 |
| 7–37 points | 32 | 2.17 | 0.68 – 3.67 | 0.007 |
| 6–36 points | 36 | 2.24 | 0.90 – 3.57 | 0.002 |
| 5–35 points | 37 | 2.22 | 0.91 – 3.52 | 0.002 |
| 5–34 points | 37 | 2.22 | 0.91 – 3.52 | 0.002 |
| 5–33 points | 37 | 2.22 | 0.91 – 3.52 | 0.002 |
| 5–32 points | 36 | 2.43 | 1.13 – 3.74 | 0.001 |
| 5–31 points | 33 | 2.87 | 1.66 – 4.11 | <0.001 |
| 5–30 points | 33 | 2.87 | 1.66 – 4.11 | <0.001 |

To estimate MCID based on the relative change in ATEC total score, the mean change in CARS2 score was calculated among participants who exhibited a 10% to 20% improvement in ATEC total score relative to their baseline score (N = 31). The estimated MCID for the CARS2 was 2.82 points (95% CI: 1.54 – 4.10; p = 0.0002). Sensitivity analysis of the MCID based on relative change in ATEC total score demonstrated excellent stability, with estimates ranging from 1.77 to 2.82 points (Table 3).

**Table 3.** Sensitivity analysis for an anchor based on the relative change in ATEC total score approach.

| Relative improvement in ATEC total score | N observations | MCID (CARS2 points) | 95% CI | P-Value |
| --- | --- | --- | --- | --- |
| 10–20% | 31 | 2.82 | 1.54 – 4.10 | <0.001 |
| 9–19% | 31 | 2.42 | 1.16 – 3.68 | <0.001 |
| 8–18% | 34 | 2.15 | 1.08 – 3.21 | <0.001 |
| **7–17%** | 38 | 2.06 | 0.96 – 3.13 | <0.001 |
| **6–16%** | 39 | 1.63 | 0.42 – 2.81 | 0.012 |
| **5–15%** | 42 | 1.77 | 0.63 – 2.87 | 0.004 |

Distribution-based approaches produced complementary estimates. One commonly used distribution-based approach estimates MCID from the baseline score variability within the study population. In this method, a change corresponding to 0.5 standard deviations (SD) of the baseline score is generally interpreted as a clinically important change, whereas 0.2 SD of the baseline score is commonly interpreted as representing a minimal clinically important change. The SD of baseline CARS2 scores in our cohort was 6.64 points. Accordingly, 0.5 SD corresponds to 3.32 points and 0.2 SD corresponds to 1.33 points.

MCID was also estimated using the Standard Error of Measurement (SEM), which incorporates both baseline score SD and test–retest reliability to account for measurement error. The SEM provides a precision-based benchmark, ensuring that the estimated MCID exceeds the inherent measurement error of the CARS2. The SEM method yielded an estimate of 1.8 points, based on a test–retest reliability coefficient of 0.9 calculated as the median value reported across the six studies summarized in Table 4.

**Table 4.** List of studies that reported test-retest reliability of CARS2.

| **Study reference** | **CARS2 test-retest reliability** | **Weeks between assessments** |
| --- | --- | --- |
| **Perry et al, (Perry et al., 2005)** | 0.77 | 12 |
| **Samadi et al, 2025 (Samadi et al., 2025)** | 0.98 | 3 |
| **Akoury-Dirani et al, 2013 (Akoury-Dirani et al., 2013)** | 0.89 | 4 |
| **Gassaloğlu et al, 2016 (İncekaş Gassaloğlu et al., 2016)** | 0.98 | 12 |
| **Pereira et al, 2008 (Pereira et al., 2008)** | 0.9 | 4 |
| **Shin et al, 1998 (Shin & Kim, 1998)** | 0.91 | 24-40 |
| **Median** | 0.9 | 4 |

## Discussion

In randomized controlled trials (RCTs), a non-significant difference between the control and treatment groups indicates insufficient statistical power due to limited sample size. However, even a statistically significant between-group difference does not necessarily imply clinical significance. To establish clinical significance, researchers rely on the group-level construct of the Minimal Clinically Important Difference (MCID), defined as the smallest between-group difference considered clinically meaningful. Accordingly, a successful trial is characterized by both statistical significance and a between-group difference that exceeds the MCID.

To our knowledge, the group-level MCID has never been empirically established for the most commonly used ASD severity measure, the *Childhood Autism Rating Scale, Second Edition* (CARS2). In this study, we addressed this gap by empirically estimating the MCID using two anchor-based and two distribution-based methods.

An anchor-based approach estimates MCID using a Patient Impression of Change (PIC) to capture perceived treatment benefit. Compared with traditional single-item Likert-scale PICs (e.g., “improved,” “slightly improved,” “no change,” “slightly worsened,” “worsened”), multi-item PICs provide greater precision and reliability, by integrating changes across multiple domains (Baker et al., 2024; Challoumas et al., 2026; Charlton et al., 2025; Zhang et al., 2023a). Additionally, single-item PICs may have limited utility in children with ASD because caregiver ratings are strongly concentrated in the direction of improvement (see Methods). Accordingly, this study used the parent-reported *Autism Treatment Evaluation Checklist* (ATEC) as a multi-item PIC. The validated 77-item ATEC assesses changes in language, cognition, sociability, and health-related behaviors. Total ATEC scores (range 0–179) were collected concurrently with CARS2 assessments every six months, and all paired CARS2/ATEC observations (N = 168) were pooled for MCID analysis.

Once a correlation of 0.41 between changes in CARS2 and absolute changes in ATEC total scores was established—exceeding the commonly accepted minimum threshold of 0.3 (Zhang et al., 2023b)—the MCID was calculated as the mean change in CARS2 score among participants who exhibited a 10- to 40-point improvement in ATEC total score, yielding an MCID of 2.39 points (p = 0.019; Table 1).

Additionally, MCID was estimated based on the relative change in ATEC total score. After establishing a correlation of 0.44 between changes in CARS2 and relative changes in ATEC total scores, the MCID was calculated as the mean change in CARS2 score among participants who exhibited a 10% to 20% improvement in ATEC total score relative to their baseline score. The estimated MCID using this method was 2.82 points (p = 0.0002).

Distribution-based approaches yielded complementary estimates of the MCID. The SD-based method produced an estimate ranging from 1.3–3.3 points, reflecting variability within the sample, whereas the SEM-based method—incorporating measurement precision and assuming a test–retest reliability of 0.9 derived from prior studies (Table 4)—generated an estimate of 1.8 points.

These results suggest that the group-level construct MCID lies within a range of 1.3–3.3 points, with the most precise estimates provided by the two anchor-based methods yielding values of 2.39–2.82 points (mean of approximately 2.6 points). Accordingly, a between-group difference of approximately 2.6 CARS2 points may be considered clinically meaningful. This estimate has direct implications for clinical trial design, including effect-size interpretation and sample size calculations.

### Comparing group-level threshold MCID to individual-level threshold MDC95

In addition to group-level evaluation, RCTs often include individual-level analyses to estimate the proportion of participants who *respond to treatment* (i.e., “responder analyses”). Responder analyses depend on an individual-level threshold that accounts for measurement error and determines whether an observed change in an individual exceeds what could be attributed to random variability. This is commonly operationalized using the Minimal Detectable Change, defined as the smallest change in score that can be interpreted as real with 95% confidence (MDC95). MDC95 is estimated from the standard error of measurement (SEM), derived from pre-treatment test–retest reliability, and is calculated as: MDC95 = 1.96 × √2 × SD × √(1-*r*), where SD denotes the standard deviation and *r* the test–retest reliability coefficient. Substituting the SD of 6.6 observed in our cohort and *r* of 0.9 (calculated as the median value reported across the six studies summarized in Table 4), we calculated an MDC95 of 5.8, which falls within the range of responder-level thresholds reported in several prior studies, with lower bounds ranging from 4.1 (Coniglio et al., 2001) to 4.5 (Jurek et al., 2022) and upper bounds ranging from 6 (Chez et al., 2000; Lemonnier et al., 2017) to 7.5 (Nagaraj et al., 2006).

As expected, the group-level construct MCID (2.6 CARS2 points) is smaller than the individual-level construct MDC95 (approximately 5.8 CARS2 points). MCID evaluates clinically meaningful differences between groups, whereas MDC95 identifies true individual-level changes beyond measurement error. Because random error is reduced when averaging across groups, MCID is typically smaller than MDC95. For example, similar relationships between MCID and MDC95 have been previously reported for measures of global cognition, such as the MoCA. In patients with dementia, the MCID has been estimated at 2 points (Krishnan et al., 2017), whereas the corresponding individual-level MDC95 at 4 points (Feeney et al., 2016). In post-stroke populations, the MCID has been estimated at 1.6–2.0 points, whereas the MDC95 has been estimated at 5.1 points (Lindvall et al., 2024).

### Limitations

A potential limitation is the use of the ATEC as an external anchor. Although parent-reported ATEC is less commonly used in clinical trials compared to clinician-rated instruments, it is particularly well suited to estimating the MCID. The MCID is defined in terms of changes perceived as meaningful to patients (or, in pediatric populations, their caregivers), and thus a caregiver-reported measure provides a direct assessment of perceived improvement in the child’s symptoms. In this context, ATEC offers an advantage over clinician-only anchors by capturing caregiver-observed changes across naturalistic settings. While parent-reported measures may introduce variability related to subjective perception, this subjectivity is intrinsic to the MCID construct itself. Therefore, the use of ATEC as an anchor represents a conceptually appropriate choice for estimating MCID.

### Clinical implications

The present findings provide an empirically grounded framework for interpreting group-level changes in CARS2 scores in ASD clinical trials. Using two anchor-based approaches, the MCID for CARS2 was estimated at 2.39–2.82 points, while distribution-based methods yielded estimates ranging from 1.3–3.3 points. The anchor-based MCID estimates fell within the range identified by the distribution-based methods, supporting the statistical reliability and clinical plausibility of the proposed MCID for CARS2. Accordingly, a between-group difference of approximately 2.6 CARS2 points may be considered clinically meaningful. This has direct implications for trial design, including sample size calculations. At the individual level, however, larger changes are required to confidently distinguish true improvement from measurement variability, potentially using an individual-level threshold of as high as 6 points. Together, these results enable more precise and transparent interpretation of CARS2 outcomes, facilitating better alignment between statistical findings, clinical relevance, and decision-making in ASD intervention research and practice.

## Acknowledgments

We sincerely thank all participants and their parents for their involvement in this study. The authors are deeply grateful to Dr. Petr Ilyinskii for his meticulous editing of the manuscript.

## Author contributions

AV designed the study. ASF, ASSS, LEPP, and EF collected data. AV and EK analyzed the data. AV wrote the paper. All authors edited the paper.

## Informed Consent

Caregivers have provided informed consent to anonymized data analysis and publication of the results. The study was conducted in compliance with the Declaration of Helsinki (World Medical Association, 2013).

## Compliance with Ethical Standards

The study protocol received IRB approval from Centro Universitário Dinâmica das Cataratas (Foz do Iguaçu, Brazil).

## Data Availability

De-identified raw data from this manuscript are available from the corresponding author upon reasonable request.

## Code availability statement

Code is available from the corresponding author upon reasonable request.

