## Supplemental material for "Identification of the Minimal Clinically Important Difference (MCID) for Childhood Autism Rating Scale Second Edition (CARS2) in children with ASD"

### Supplementary Material

Supplementary Table 1: *Childhood Autism Rating Scale Second Edition* (CARS2) [1] is a 15-item behavioral rating scale developed to quantitatively assess the severity of ASD. CARS2 works by rating a child's behavior, characteristics, and abilities against the expected developmental growth of a typical child. Each item is scored from 1 to 4, with a score of 1 indicating behavior typical for a child's age; 2, mildly abnormal; 3, moderately abnormal; and 4, severely abnormal. CARS2 total score ranges from 15 (no evidence of autism) to 60 (severe ASD), with 30 being the cutoff rate for a diagnosis of mild autism. Scores 30-37 indicate mild to moderate autism, while scores between 38 and 60 are characterized as severe autism [2].

| |
|---|
| 1. Relating to people |
| 2. Imitation (how child imitated verbal and nonverbal acts) |
| 3. Emotional Response |
| 4. Body Use |
| 5. Object Use (interest in toys and how he is using them) |
| 6. Adaption to Change |
| 7. Visual Response (rating of unusual visual attention patterns when required to look at objects, e.g. avoiding eye contact) |
| 8. Listening Response |
| 9. Taste, Smell, and Touch Response and Use |
| 10. Fear or nervousness |
| 11. Verbal communication |
| 12. Nonverbal Communication |
| 13. Activity level |
| 14. Level and consistency of intellectual response |
| 15. General Impressions |

Supplementary Table 2: *Autism Treatment Evaluation Checklist* (ATEC) [3] comprises four subscales: 1) Speech/Language/Communication, 2) Sociability, 3) Sensory/Cognitive Awareness, and 4) Physical/Health/Behavior. ATEC total score ranges from 0 to 179.
While the CARS2 is specifically designed to diagnose and categorize the severity of autism, the ATEC was actually designed by the Autism Research Institute (ARI) for a different purpose: to measure the effectiveness of treatments over time. Because of this, the ATEC is not officially a diagnostic tool.

ATEC Subscale 1: Speech/Language/Communication. The answers choices were: not true, somewhat true, very true.

| |
|---|
| 1. Knows own name |
| 2. Responds to 'No' or 'Stop' |
| 3. Can follow some commands |
| 4. Can use one word at a time (No!, Eat, Water, etc.) |
| 5. Can use 2 words at a time (Don't want, Go home) |
| 6. Can use 3 words at a time (Want more milk) |
| 7. Knows 10 or more words |
| 8. Can use sentences with 4 or more words |
| 9. Explains what he/she wants |
| 10. Asks meaningful questions |
| 11. Speech tends to be meaningful/relevant |
| 12. Often uses several successive sentences |
| 13. Carries on fairly good conversation |
| 14. Has normal ability to communicate for his/her age |

ATEC subscale 2: Sociability**.** The answers choices were: not true, somewhat true, very true.

| |
|---|
| 1. Seems to be in a shell – you cannot reach him/her |
| 2. Ignores other people |
| 3. Pays little or no attention when addressed |
| 4. Uncooperative and resistant |
| 5. No eye contact |
| 6. Prefers to be left alone |
| 7. Shows no affection |
| 8. Fails to greet parents |
| 9. Avoids contact with others |
| 10. Does not imitate |
| 11. Dislikes being held/cuddled |
| 12. Does not share or show |
| 13. Does not wave 'bye bye' |
| 14. Disagreeable/not compliant |
| 15. Temper tantrums |
| 16. Lacks friends/companions |
| 17. Rarely smiles |
| 18. Insensitive to other's feelings |
| 19. Indifferent to being liked |
| 20. Indifferent if parent(s) leave |

ATEC subscale 3: Sensory/Cognitive awareness**.** The answers choices were: not true, somewhat true, very true.

| |
|---|
| 1. Responds to own name |
| 2. Responds to praise |
| 3. Looks at people and animals |
| 4. Looks at pictures (and T.V.) |
| 5. Does drawing, coloring, art |
| 6. Plays with toys appropriately |
| 7. Appropriate facial expression |
| 8. Understands stories on T.V. |
| 9. Understands explanations |
| 10. Aware of environment |
| 11. Aware of danger |
| 12. Shows imagination |
| 13. Initiates activities |
| 14. Dresses self |
| 15. Curious, interested |
| 16. Venturesome - explores |
| 17. "Tuned in" — Not spacey |
| 18. Looks where others are looking |

ATEC subscale 4: Health/Physical/Behavior**.** The answers choices were: not a problem, minor problem, moderate problem, and serious problem.

| |
|---|
| 1. Bed-wetting |
| 2. Wets pants/diapers |
| 3. Soils pants/diapers |
| 4. Diarrhea |
| 5. Constipation |
| 6. Sleep problems |
| 7. Eats too much/too little |
| 8. Extremely limited diet |
| 9. Hyperactive |
| 10. Lethargic |
| 11. Hits or injures self |
| 12. Hits or injures others |
| 13. Destructive |
| 14. Sound-sensitive |
| 15. Anxious/fearful |
| 16. Unhappy/crying |
| 17. Seizures |
| 18. Obsessive speech |
| 19. Rigid routines |
| 20. Shouts or screams |
| 21. Demands sameness |
| 22. Often agitated |
| 23. Not sensitive to pain |

| |
|---|
| 24. “Hooked” or fixated on certain objects/topics |
| 25. Repetitive movements (stimming, rocking, etc.) |

**Supplementary Material References**

1. Schopler, E., Reichler, R. J. & Renner, B. R. *The Childhood Autism Rating Scale (CARS)*. (Western Psychological Services Los Angeles, CA, 2002).
2. Schopler, E., Reichler, R. J., DeVellis, R. F. & Daly, K. Toward objective classification of childhood autism: Childhood Autism Rating Scale (CARS). *J. Autism Dev. Disord.* (1980).
3. Rimland, B. & Edelson, S. M. Autism treatment evaluation checklist (ATEC). *Autism Res. Inst. San Diego CA* http://www.autism.com (1999).
